# From Clinical Judgment to Large Language Models: Benchmarking Predictive Approaches for Unplanned Hospital Admissions

**DOI:** 10.1101/2025.09.09.25335411

**Authors:** Bernardo Neves, Mário J. Silva

## Abstract

**Background:** While machine learning (ML) models show strong performance for predicting unplanned hospital visits, their clinical utility relative to physician judgment remains unclear. Large language models (LLMs) offer a promising middle ground, potentially combining algorithmic accuracy with human-interpretable reasoning.

**Objective:** To directly compare the predictive performance of physicians, structured ML models, and LLMs for forecasting 30-day emergency department (ED) visits and unplanned hospital admissions under equivalent data conditions.

**Methods:** We selected 404 cases from structured EHR data and converted them into synthetic clinical vignettes using GPT-5. Thirty-five physicians evaluated these vignettes, while CLMBR-T (a machine learning model trained on structured EHR data) was applied to the original data. Eight LLMs evaluated the same vignettes. We compared discriminative performance (AUROC, AUPRC), calibration (Brier score, Expected Calibration Error), and confidence-performance relationships across all methods.

**Results:** CLMBR-T achieved the highest discriminative performance (AUROC 0.79, 95% CI: 0.75-0.83; AUPRC 0.78, 95% CI: 0.72-0.83), followed by large LLMs (DeepSeek V3, Claude 4.1 Opus, GPT-5; AUROC 0.74). Pooled physicians performed lowest (AUROC 0.65, 95% CI: 0.59-0.70; AUPRC 0.61, 95% CI: 0.54-0.68). However, LLMs showed stronger alignment with physician reasoning (correlation r=0.51-0.65) compared to CLMBR-T (r=0.37). CLMBR-T demonstrated superior confidence calibration with significant confidence-performance correlation (r=0.21, p<0.001), while physicians showed poor calibration (r=0.07, p=0.17). Individual physician performance varied widely (AUROC 0.55-0.83), with three out of 35 physicians exceeding the ML benchmark.

**Conclusions:** ML models trained on structured EHR data outperform both physicians and LLMs in predictive accuracy and confidence calibration, though LLMs achieved competitive zero-shot performance and better approximated human clinical reasoning. These findings suggest hybrid approaches combining high-performance ML screening with interpretable LLM explanations may optimize both accuracy and clinical adoption. The substantial variability in physician performance highlights limitations of benchmarking against “average” clinical judgment.

## 1 Introduction

Unplanned hospital admissions represent a substantial burden for patients and healthcare systems worldwide, with emergency department visits and short-term readmissions often considered potentially preventable through targeted interventions [1]. This challenge has driven decades of research into predictive modeling approaches using administrative and electronic health record data to identify high-risk patients and enable proactive care management.

Traditional risk prediction models evolved from simple rule-based scores such as the LACE index and HOSPITAL score, which achieved modest performance with AUROCs of approximately 0.65-0.75 [2]. The adoption of machine learning methods using electronic health record data has substantially improved predictive accuracy, with ensemble approaches such as gradient boosting and random forests consistently achieving AUROCs between 0.70 and 0.85 [3]. However, these gains in algorithmic performance raise fundamental questions about clinical utility, as the opacity of machine learning models often limits trust and adoption in clinical settings.

Physicians’ own prognostic judgment provides a natural benchmark for evaluating predictive models. Research on clinical forecasting ability reveals that while physician estimates often correlate with actual outcomes, they are frequently inaccurate and exhibit systematic optimism bias [4]. Studies of unplanned hospital visit prediction specifically show moderate physician discriminative performance, with AUROCs typically ranging from 0.57 to 0.73 across different care settings [5]. These findings suggest significant room for improvement through algorithmic support, yet direct comparisons between physician judgment and machine learning models remain limited and have yielded mixed results.

Large language models have recently emerged as a potentially transformative approach that may bridge the gap between high-performing but opaque machine learning systems and interpretable human reasoning. Early clinical applications demonstrate promising performance, with models such as NYUTron achieving superior readmission prediction compared to regression baselines and GPT-4 showing physician-comparable accuracy in emergency department triage tasks [6]. Unlike traditional machine learning approaches, large language models can provide human-interpretable rationales for their predictions, potentially combining algorithmic accuracy with clinical reasoning transparency.

Despite these advances, no study has directly compared the predictive performance of physicians, structured machine learning models, and large language models under equivalent data conditions for forecasting unplanned hospital visits. This gap leaves critical questions unanswered about the relative strengths of each approach in terms of discriminative accuracy, probability calibration, and alignment with clinical reasoning patterns. Understanding these trade-offs is essential for developing effective clinical decision support systems that optimize both predictive performance and clinical adoption.

This study addresses this knowledge gap by conducting a systematic head-to-head comparison of physicians, a state-of-the-art machine learning model trained on structured electronic health record data, and multiple large language models in predicting 30-day emergency department visits and unplanned hospital admissions. Through careful experimental design using synthetic clinical vignettes generated from real patient data, we evaluate not only discriminative performance but also calibration quality, confidence relationships, and alignment with human clinical reasoning to inform the development of hybrid approaches that leverage the complementary strengths of each method.

## 2. Methods

### 2.1 Overview of Study Design

We compared the predictive performance of three approaches—structured EHR-based machine learning, physicians, and large language models (LLMs)—for forecasting 30-day emergency department (ED) visits and unplanned hospital admissions. As illustrated in the graphical abstract (Figure 1), the study workflow began with structured EHR data from Hospital da Luz Lisboa stored in OMOP-CDM format. A logistic regression model trained on pretrained CLMBR-T patient representations [7] was used as the structured ML baseline. From the held-out test set, a random sample of cases was selected and converted into synthetic clinical vignettes using an LLM. These vignettes were independently evaluated by physicians and by several general-purpose LLMs. We then compared predictions across all methods in terms of discrimination, calibration, classification metrics, and confidence-performance relationships.

**Figure 1.**
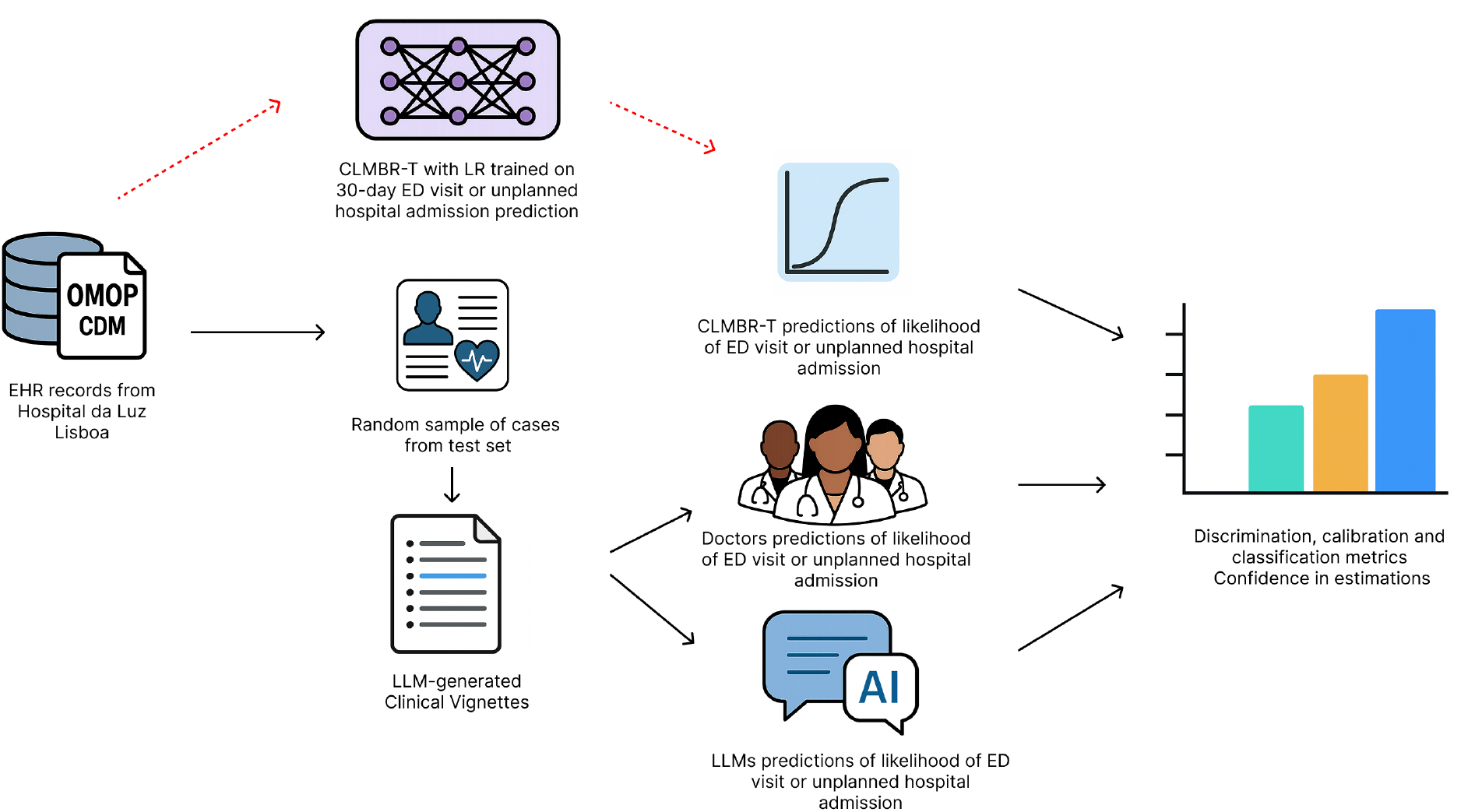
Graphical abstract of the study workflow: structured EHR data were used to train a logistic regression model on pretrained CLMBR-T representations; a sample of test cases was transformed into LLM-generated clinical vignettes; physicians and LLMs provided predictions of 30-day ED visits or unplanned admissions; performance was compared across methods in terms of discrimination, calibration, and confidence in estimations.

### 2.2 Machine Learning Model

For the structured EHR–based model, we used a logistic regression classifier trained on fixed patient representations generated by a pretrained CLMBR–T model [7]. In prior benchmarking on our cohort, this approach achieved the strongest performance, so we adopted it as our structured-data baseline. The classifier was trained to predict 30-day ED visits and unplanned hospital admissions from longitudinal clinical trajectories of 100,000 patients, and evaluated on a held-out test set of 50,000 patients. From this test set, we sampled cases to enable head-to-head comparisons with LLMs and physicians (details below).

### 2.3 Patient Selection and Eligibility

From the held-out test set used in the ML model development, we selected 404 patients meeting the following criteria:

- Evidence of multiple chronic conditions at any time in their clinical record;
- Sufficient data history (excluded lowest quartile by number of OMOP codes recorded prior to prediction point);
- Random selection of one visit per patient as the prediction point (discharge for inpatient episodes, visit time for outpatient/ED visits).

The prediction task was to forecast whether an ED visit or unplanned hospitalization would occur within 30 days of the prediction point.

### 2.4 Conversion to Clinical Vignettes

We developed a systematic pipeline to convert structured OMOP data into clinical narratives:

1. **Data Extraction:** All clinical events prior to the prediction point were extracted, including demographics, visit types, diagnoses, laboratory results, procedures, and medications.
2. **Text Generation:** A rule-based Python script converted structured codes to natural language using OHDSI vocabularies, organizing information by clinical category and temporal relevance.
3. **Narrative Synthesis:** GPT-5 transformed the structured text into professional clinical vignettes with three standardized sections:
  - Current Visit: Age, sex, setting, presentation, tests/treatments
  - Past Year: Recent visits in reverse chronological order
  - Remote History: Earlier findings organized by category

The prompt for vignette generation was iteratively refined and validated by consensus among six physicians for clinical realism and clarity.

### 2.5 Physician Evaluation

We recruited 35 practicing physicians from HLL through institutional networks. Each physician evaluated 24 vignettes (balanced 50% outcome prevalence) including 4 anchor cases for reliability assessment and 20 additional cases. For each vignette, physicians provided:

- Likelihood rating (1-5 Likert scale) for 30-day ED visit or unplanned admission
- Confidence rating (1-5 Likert scale) in their assessment

Participants were stratified by specialty, experience level, and ED work exposure. The study received local Ethics Comitee approval (ID 804, CES 33/2025).

### 2.6 Large Language Model Evaluation

Eight LLMs evaluated the clinical vignettes using standardized prompts requesting:

- Likelihood rating (1-5 Likert scale)
- Confidence rating (1-5 Likert scale)
- Predicted probability (0-1)

Evaluated models included:

- **Proprietary:** GPT-5, Claude 4.1 Opus, GPT-4o
- **Open-source:** DeepSeek V3, GPT-oss 120B, Gemma 3 120B, Qwen 3 32B, Qwen 4B

All models used identical configurations (temperature=0.1, max tokens=100) with structured JSON output format.

### 2.7 Statistical Analysis

We assessed discriminative performance using AUROC and AUPRC, calibration using Brier score and Expected Calibration Error (ECE), and classification metrics at Youden-optimal thresholds. Statistical inference used study-aware bootstrap resampling (n=1,000) preserving the clustered experimental design, in order to account to repeated visits from the same patient.

Confidence analysis examined correlations between self-reported confidence and accuracy. For ML models, we converted Shannon entropy to a 1-5 confidence scale for comparison. Inter-rater reliability was assessed using Krippendorff’s alpha for anchor questions and Cohen’s weighted kappa for paired evaluations.

Human-AI alignment was measured through probability correlations between physician predictions and model outputs, providing insight into reasoning similarity.

## 3 Results

### 3.1 Participants description and inter-rater reliability analysis

Thirty-five physicians completed all 24 clinical vignettes, with participants attributes summarized in Table 1. The sample was predominantly female (66%) and specialized in Internal Medicine (60%), with diverse clinical experience ranging from less than 5 to more than 20 years of practice. Most participants (71%) reported to work regularly in the ED.

**Table 1:**
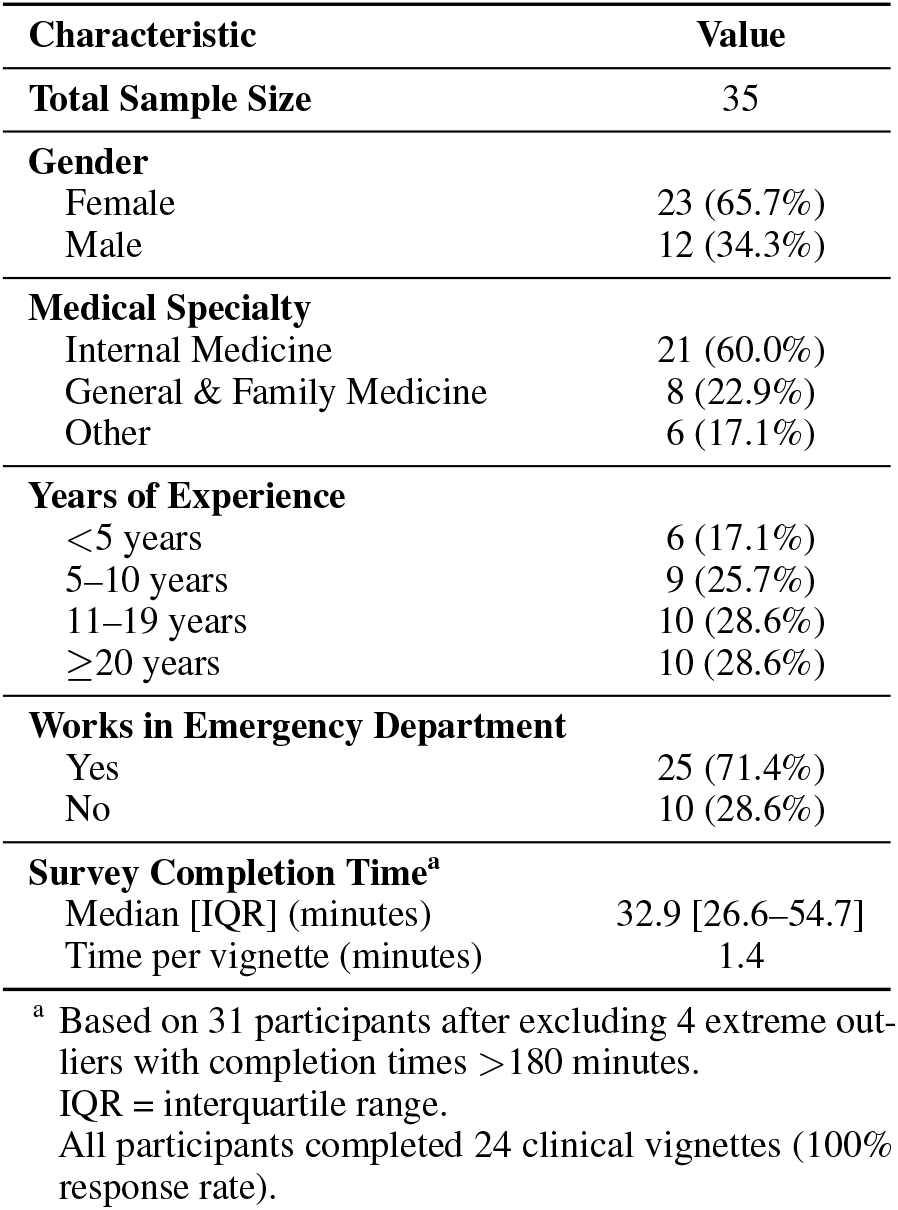
Participant characteristics.

| Characteristic | Value |
| --- | --- |
| **Total Sample Size** | 35 |
| **Gender** |  |
| Female | 23 (65.7%) |
| Male | 12 (34.3%) |
| **Medical Specialty** |  |
| Internal Medicine | 21 (60.0%) |
| General & Family Medicine | 8 (22.9%) |
| Other | 6 (17.1%) |
| **Years of Experience** |  |
| <5 years | 6 (17.1%) |
| 5–10 years | 9 (25.7%) |
| 11–19 years | 10 (28.6%) |
| ≥20 years | 10 (28.6%) |
| **Works in Emergency Department** |  |
| Yes | 25 (71.4%) |
| No | 10 (28.6%) |
| **Survey Completion Time<sup>a</sup>** |  |
| Median [IQR] (minutes) | 32.9 [26.6–54.7] |
| Time per vignette (minutes) | 1.4 |

<sup>a</sup> Based on 31 participants after excluding 4 extreme outliers with completion times >180 minutes.
IQR = interquartile range.
All participants completed 24 clinical vignettes (100% response rate).

A total of 840 assessments were performed by participants, from a a total of 404 unique cases (2.1 evaluations per case on average). Median time to completion of the 24 cases was 36 minutes (IQR 27-55), indicating appropriate task duration and participant engagement.

As shown in Table 2, inter-rater reliability on the four anchor questions was moderate, with Krippendorff’s *α* = 0.65 (95% CI: 0.57–0.76). In contrast, agreement on the paired questions was poor, with a mean weighted Cohen’s *κ* of 0.11 (range: *−* 0.33 to 0.78) and only 19.2% exact agreement, although within-one-level agreement reached 62.3%. Reliability for self-reported confidence ratings was low (Krippendorff’s *α* = 0.35), indicating that certainty scores were largely idiosyncratic. Subgroup analyses showed higher consistency among family medicine doctors (Krippendorff’s *α* = 0.88) and among mid-career physicians with 5–10 years of experience (Krippendorff’s *α* = 0.86), whereas both junior doctors (*<*5 years, *α* = 0.51) and senior doctors (≥20 years, α = 0.55) demonstrated lower reliability.

**Table 2:**
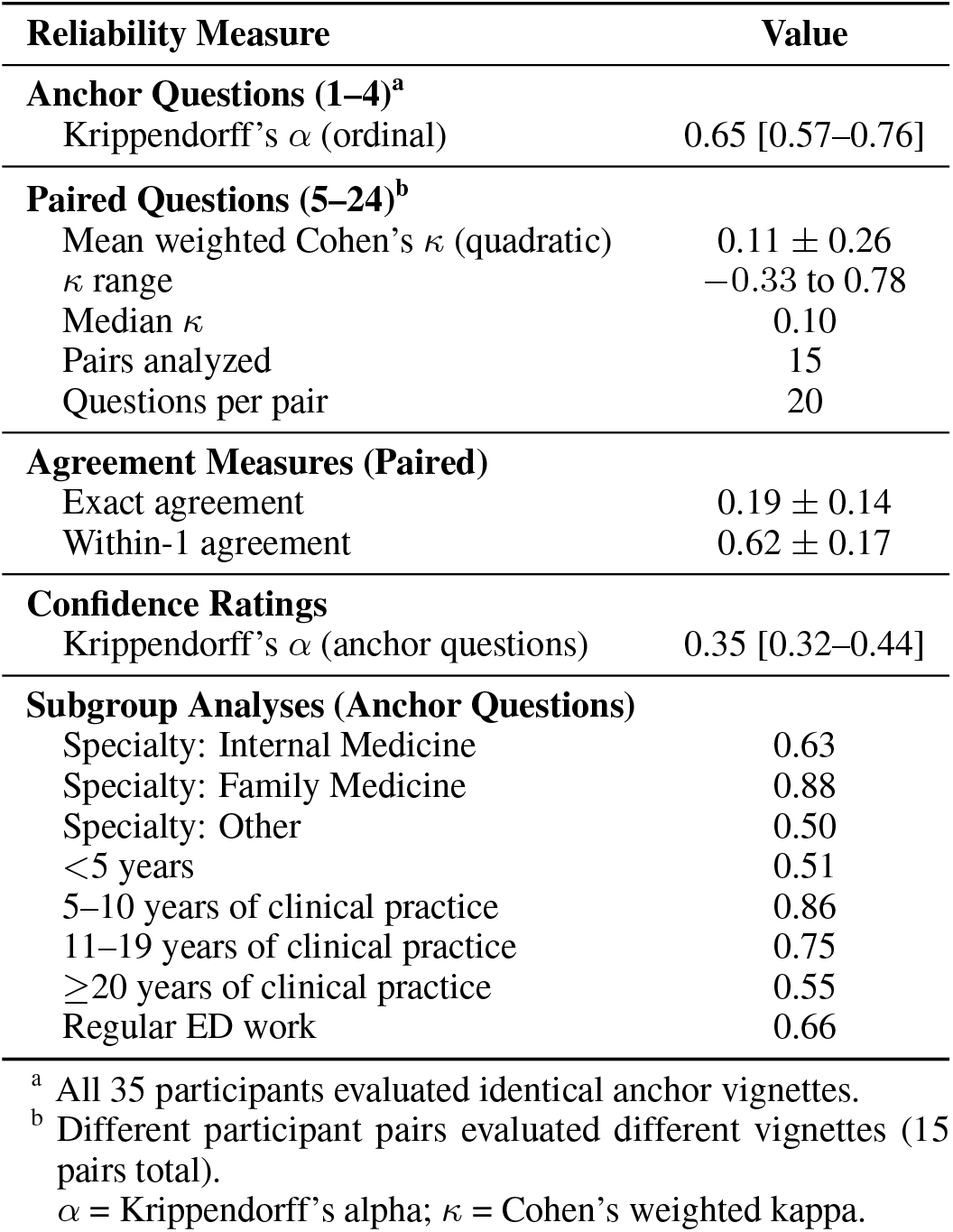
Inter-rater reliability results, overall and by subgroups.

| Reliability Measure | Value |
| --- | --- |
| **Anchor Questions (1–4)<sup>a</sup>** |  |
| Krippendorff’s $\alpha$ (ordinal) | 0.65 [0.57–0.76] |
| **Paired Questions (5–24)<sup>b</sup>** |  |
| Mean weighted Cohen’s $\kappa$ (quadratic) | $0.11 \pm 0.26$ |
| $\kappa$ range | –0.33 to 0.78 |
| Median $\kappa$ | 0.10 |
| Pairs analyzed | 15 |
| Questions per pair | 20 |
| **Agreement Measures (Paired)** |  |
| Exact agreement | $0.19 \pm 0.14$ |
| Within-1 agreement | $0.62 \pm 0.17$ |
| **Confidence Ratings** |  |
| Krippendorff’s $\alpha$ (anchor questions) | 0.35 [0.32–0.44] |
| **Subgroup Analyses (Anchor Questions)** |  |
| Specialty: Internal Medicine | 0.63 |
| Specialty: Family Medicine | 0.88 |
| Specialty: Other | 0.50 |
| <5 years | 0.51 |
| 5–10 years of clinical practice | 0.86 |
| 11–19 years of clinical practice | 0.75 |
| $\geq 20$ years of clinical practice | 0.55 |
| Regular ED work | 0.66 |

<sup>a</sup> All 35 participants evaluated identical anchor vignettes.<sup>b</sup> Different participant pairs evaluated different vignettes (15 pairs total). $\alpha$ = Krippendorff’s alpha; $\kappa$ = Cohen’s weighted kappa.

### 3.2 Discriminative performance comparison

#### Overall performance

Across all 404 cases, discrimination differed markedly between doctors, CLMBR-T, and LLMs. As summarized in Table 3 and visualized in Figures 2 and 3, CLMBR-T achieved the highest performance (AUROC 0.79, 95% CI: 0.75–0.83; AUPRC 0.78, 95% CI: 0.72–0.83), followed by the best LLMs (DeepSeek, Claude, GPT-5; AUROC *∼* 0.74, AUPRC *∼* 0.70–0.71). Pooled physicians performed lowest overall (AUROC 0.65, 95% CI: 0.59–0.70; AUPRC 0.61, 95% CI: 0.54–0.68), establishing a clear hierarchy: structured ML *>* larger LLMs *>* smaller LLMs *>* doctors.

**Table 3:**
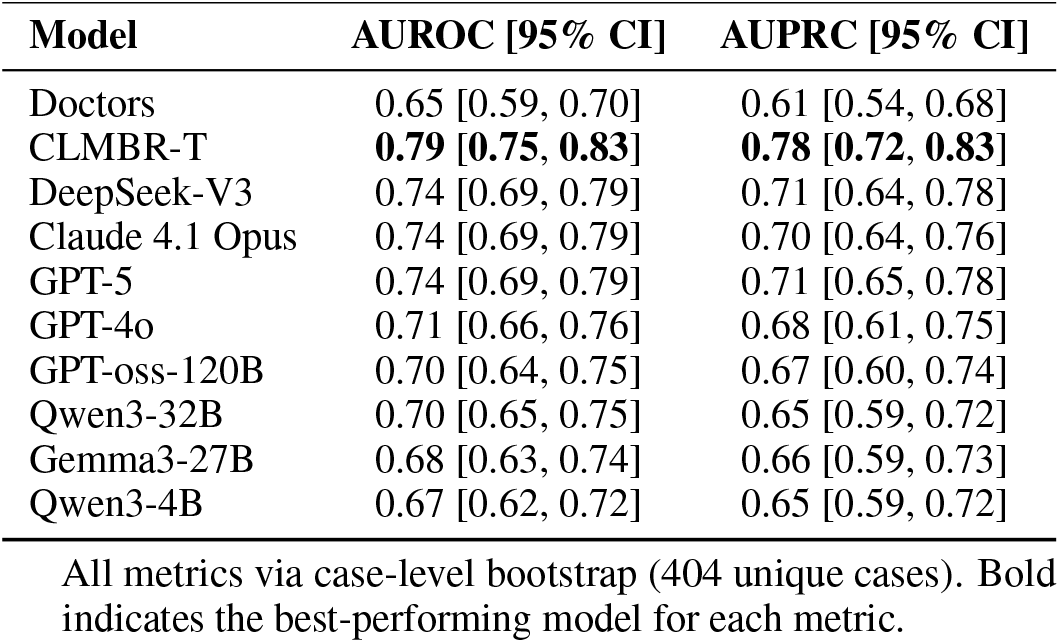
Overall discrimination of doctors and all methods on the 404 unique evaluated cases.

| Model | AUROC [95% CI] | AUPRC [95% CI] |
| --- | --- | --- |
| Doctors | 0.65 [0.59, 0.70] | 0.61 [0.54, 0.68] |
| CLMBR-T | **0.79 [0.75, 0.83]** | **0.78 [0.72, 0.83]** |
| DeepSeek-V3 | 0.74 [0.69, 0.79] | 0.71 [0.64, 0.78] |
| Claude 4.1 Opus | 0.74 [0.69, 0.79] | 0.70 [0.64, 0.76] |
| GPT-5 | 0.74 [0.69, 0.79] | 0.71 [0.65, 0.78] |
| GPT-4o | 0.71 [0.66, 0.76] | 0.68 [0.61, 0.75] |
| GPT-oss-120B | 0.70 [0.64, 0.75] | 0.67 [0.60, 0.74] |
| Qwen3-32B | 0.70 [0.65, 0.75] | 0.65 [0.59, 0.72] |
| Gemma3-27B | 0.68 [0.63, 0.74] | 0.66 [0.59, 0.73] |
| Qwen3-4B | 0.67 [0.62, 0.72] | 0.65 [0.59, 0.72] |

All metrics via case-level bootstrap (404 unique cases). Bold indicates the best-performing model for each metric.

**Figure 2.**
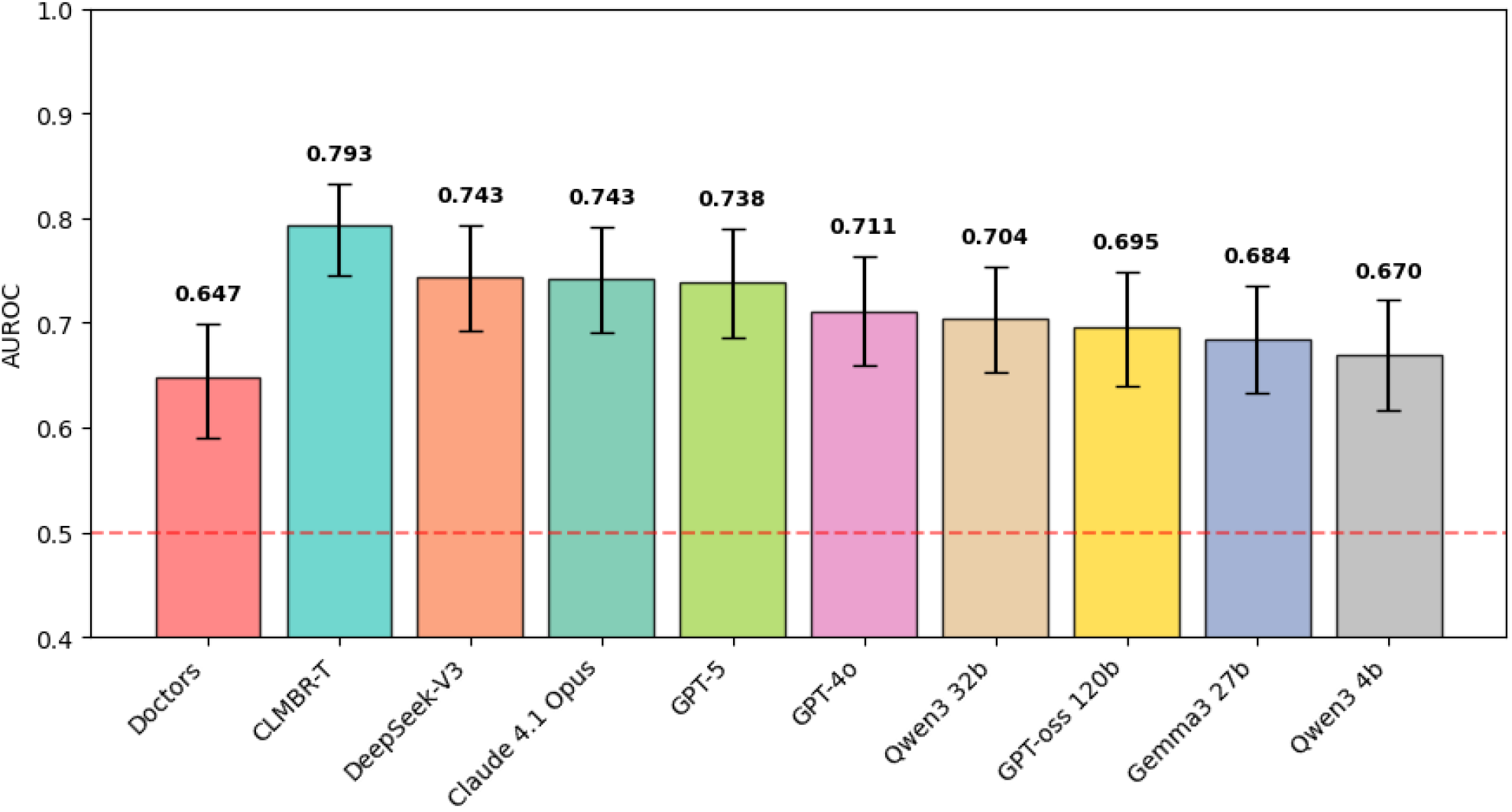
Overall discrimination performance across methods (AUROC with 95% CIs).

**Figure 3.**
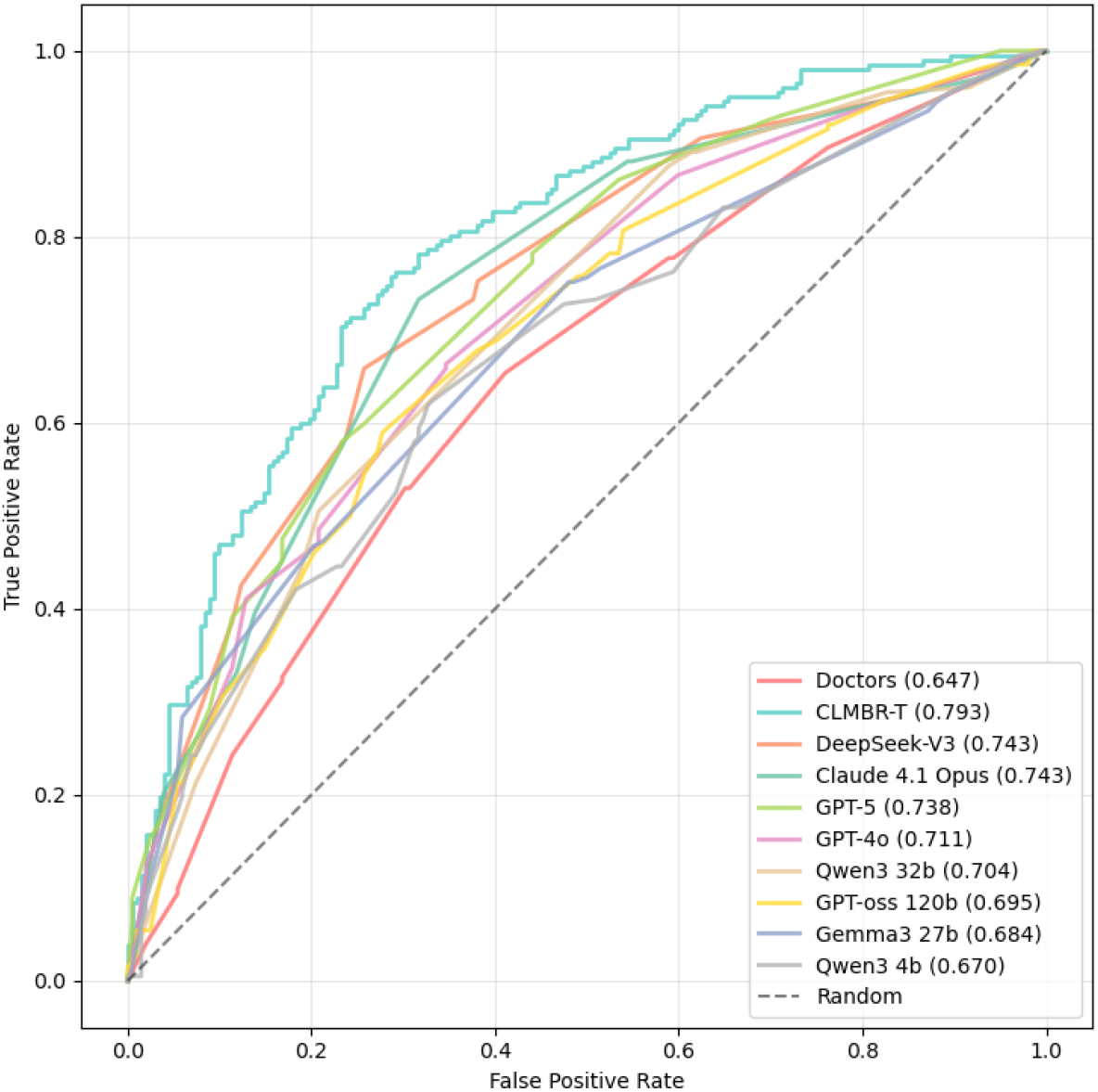
ROC curves across all evaluated methods.

#### Outcome-specific comparisons

Performance patterns were highly consistent across both endpoints—30-day ED visits and unplanned admissions. As shown in Table 4, pooled physicians showed similar discrimination for ED visits (AUROC 0.65; AUPRC 0.61) and admissions (AUROC 0.65; AUPRC 0.61). CLMBR-T remained superior for both outcomes (AUROC *≈* 0.79; AUPRC *≈* 0.78). LLM rankings were stable across endpoints (DeepSeek/Claude/GPT-5 leading; Gemma/Qwen variants with lower performance), indicating that relative ordering does not depend on outcome type.

**Table 4:** Outcome-specific AUROC values.

| Method | ED Visit | Unplanned Admission |
| --- | --- | --- |
| Doctors | 0.65 [0.58, 0.70] | 0.65 [0.59, 0.70] |
| CLMBR-T | **0.79 [0.75, 0.83]** | **0.79 [0.75, 0.83]** |
| DeepSeek-V3 | 0.74 [0.69, 0.79] | 0.74 [0.69, 0.79] |
| Claude Opus | 0.74 [0.69, 0.79] | 0.74 [0.69, 0.79] |
| GPT-5 | 0.74 [0.68, 0.78] | 0.74 [0.69, 0.78] |
| GPT-4o | 0.71 [0.65, 0.75] | 0.71 [0.66, 0.76] |
| GPT-oss-120B | 0.69 [0.64, 0.74] | 0.69 [0.64, 0.74] |
| Qwen3-32B | 0.70 [0.65, 0.75] | 0.70 [0.65, 0.75] |
| Gemma3-27B | 0.68 [0.63, 0.73] | 0.68 [0.63, 0.74] |
| Qwen3-4B | 0.67 [0.62, 0.72] | 0.67 [0.62, 0.72] |

Values rounded to two decimals. Bold indicates the best-performing model for each metric. Case-level bootstrap CIs.

#### Individual physicians and subgroups

Across the 35 participating physicians, discrimination varied widely (AUROC range: 0.55–0.830; median: 0.66). Three physicians (8.6%) exceeded the CLMBR-T benchmark (0.79), with the best individual reaching 0.83 (gap +0.04 vs. CLMBR-T). By years of experience, mean AUROC was highest for mid-career physicians (5–10 years: 0.70), lower for juniors (*<*5 years: 0.64) and seniors (*≥* 20 years: 0.68), with intermediate performance among those with 11–19 years (0.66); differences across experience groups were not statistically significant (ANOVA *p* = 0.579). Individual results are visualized in Figure 4, which overlays reference lines for CLMBR-T (0.79) and pooled doctors (0.68).

**Figure 4.**
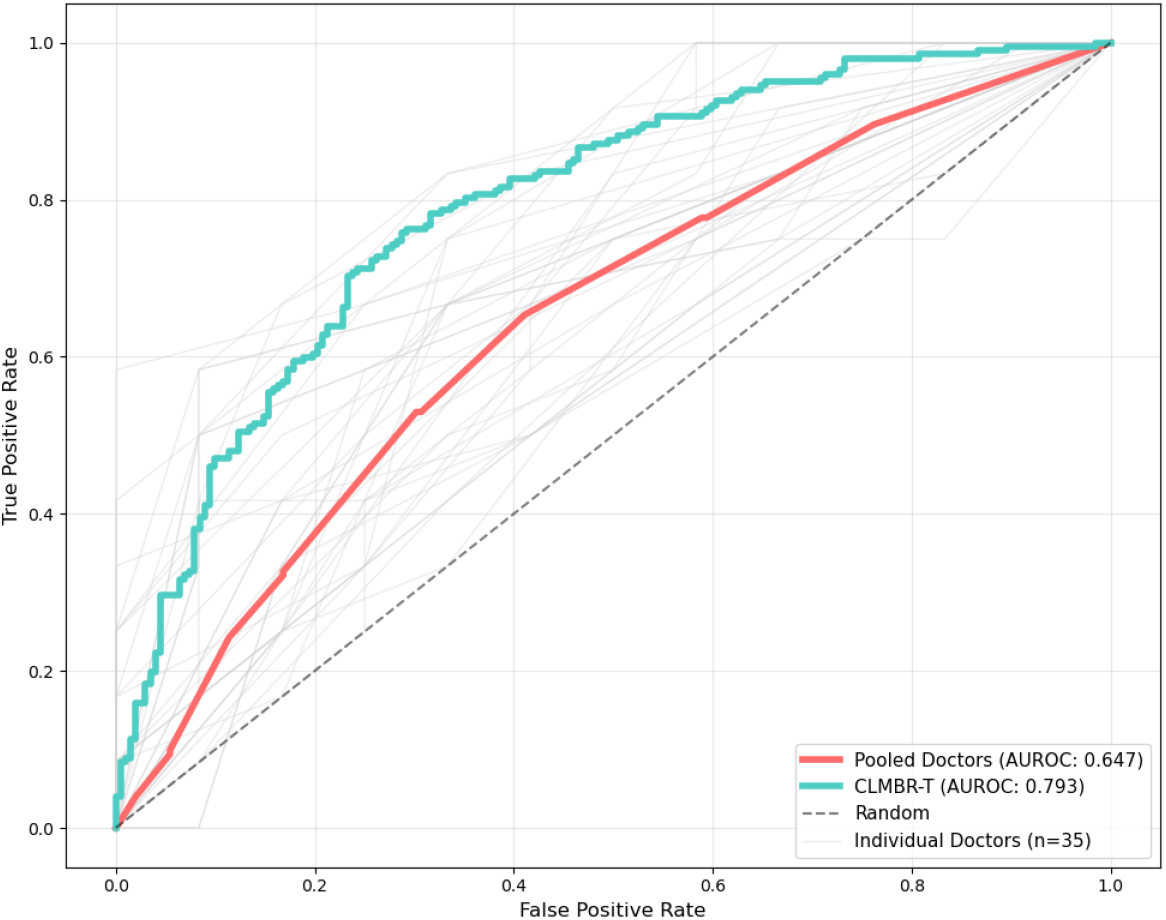
AUROC curves of individual physicians (n=35; thin lines) and the CLMBR-T model (bold line). The area under each curve is shown per physician; the dashed horizontal reference marks the pooled-doctor AUROC (0.647), and the solid reference marks CLMBR-T’s AUROC (0.793).

### 3.3 Calibration performance

As summarized in Table 5, CLMBR-T displayed the best overall calibration across the 404 cases, achieving the lowest Brier score (0.19, 95% CI: 0.17–0.22) and the lowest ECE (0.08, 95% CI: 0.06–0.13). Doctors ranked second on ECE (0.13, 95% CI: 0.09–0.18) but showed a higher Brier score (0.27, 95% CI: 0.25–0.31), indicating probabilities that are reasonably well aligned with observed frequencies yet less sharp than the ML model. Reliability diagrams are depicted in Figure 5

**Table 5:** Overall calibration across methods.

| Method | Brier Score [95% CI] | ECE [95% CI] |
| --- | --- | --- |
| Doctors | 0.27 [0.25, 0.31] | 0.13 [0.09, 0.18] |
| CLMBR-T | **0.19 [0.17, 0.22]** | **0.08 [0.06, 0.13]** |
| DeepSeek-V3 | 0.26 [0.23, 0.28] | 0.22 [0.18, 0.26] |
| Claude Opus | 0.23 [0.21, 0.26] | 0.15 [0.12, 0.20] |
| GPT-5 | 0.27 [0.24, 0.30] | 0.24 [0.20, 0.29] |
| GPT-4o | 0.27 [0.24, 0.30] | 0.22 [0.18, 0.27] |
| GPT-oss-120B | 0.29 [0.26, 0.32] | 0.26 [0.22, 0.31] |
| Qwen3-32B | 0.28 [0.26, 0.30] | 0.24 [0.20, 0.28] |
| Gemma3-27B | 0.28 [0.25, 0.30]<sup>a</sup> | 0.23 [0.19, 0.28]<sup>a</sup> |
| Qwen3-4B | 0.35 [0.31, 0.38] | 0.34 [0.30, 0.39] |

<sup>a</sup> Computed on 403 cases due to one missing prediction.
ECE = Expected Calibration Error (10-bin quantile). Bold indicates the best-performing model for each metric.

**Figure 5.**
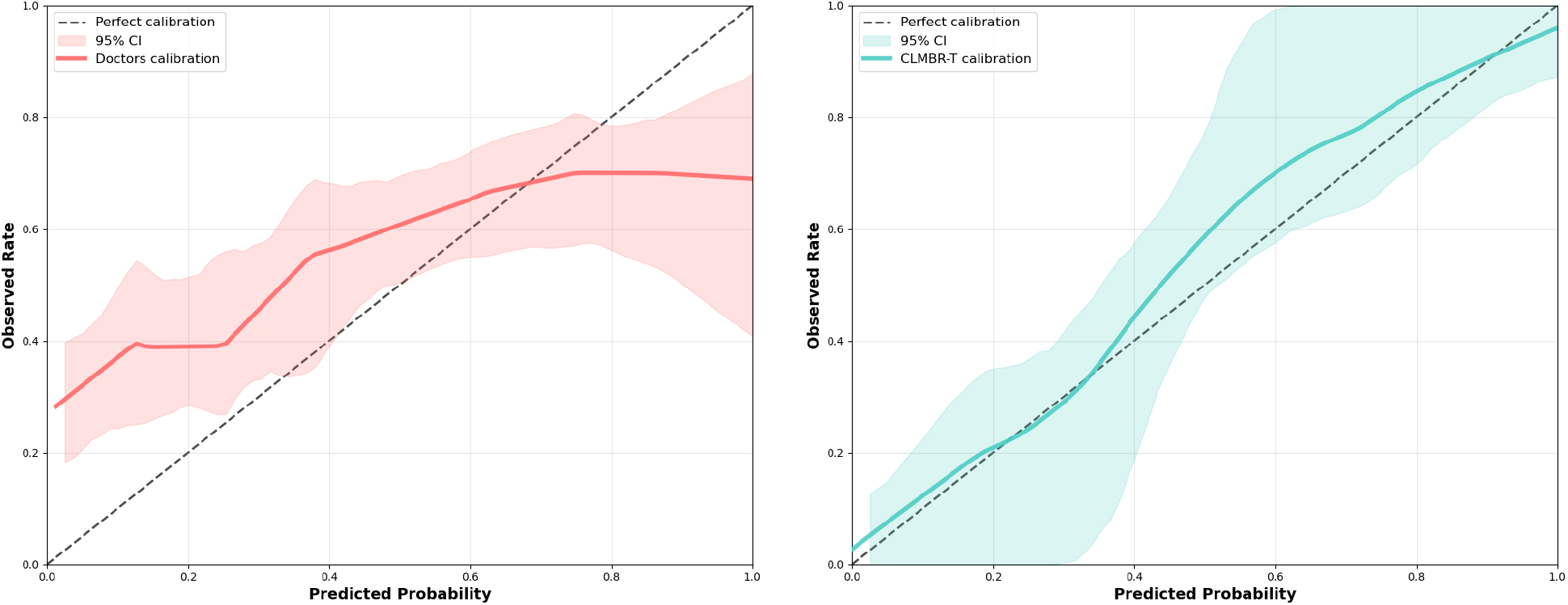
Reliability diagrams with 95% bootstrap bands for Doctors (left) and CLMBR-T (right). Lower ECE corresponds to closer adherence to the diagonal; lower Brier reflects better overall accuracy and sharpness.

Among LLMs, *Claude Opus* showed the best calibration (Brier 0.23; ECE 0.151), followed by *DeepSeek-V3*, while *Qwen3-4B* was the least calibrated (0.35; 0.34). The reliability diagrams reflect this pattern: CLMBR-T tracks the 45^*?*^ identity line most closely with narrow bootstrap bands; the pooled-physician curve is also near the identity line but with wider bands and a narrower spread of predicted risks—consistent with their relatively low ECE yet higher Brier.

### 3.4 Classification and reclassification metrics

Under thresholds selected by maximizing Youden’s index (Table 6), CLMBR-T provided the most balanced classification performance, yielding the highest F1 score (0.74; 95% CI: 0.69–0.79) and accuracy (0.74; 95% CI: 0.69–0.78), with sensitivity (0.76; 95% CI: 0.70–0.82) and specificity (0.71; 95% CI: 0.65–0.77) in close alignment. Among LLMs, Claude Opus performed best (F1 = 0.72), followed by DeepSeek-V3 (F1 = 0.69). Pooled physicians achieved an F1 of 0.63 and accuracy of 0.621, representing absolute deficits of approximately 0.11 and 0.11 relative to CLMBR-T, respectively. Operating-point characteristics differed across models: Gemma3-27B favored recall (sensitivity = 0.75) at the expense of specificity (0.52), whereas GPT-5 and Qwen3-32B adopted more conservative thresholds, producing higher specificity (0.766 and 0.792) but lower sensitivity (0.580 and 0.51). DeepSeek-V3 and GPT-4o occupied a more balanced regime, and Claude 4.1 Opus most closely approximated CLMBR-T’s balance across sensitivity, specificity, precision, and F1. These patterns were consistent across both endpoints (ED visits and unplanned admissions), with rankings and magnitudes remaining stable by outcome.

**Table 6:** Overall classification metrics at Youden-optimal thresholds.

| Method | Threshold | Accuracy | Sensitivity | Precision | F1 |
| --- | --- | --- | --- | --- | --- |
| Doctors | 0.38 | 0.62 [0.57, 0.67] | 0.65 [0.59, 0.72] | 0.61 [0.55, 0.68] | 0.63 [0.57, 0.68] |
| CLMBR-T | 0.42 | **0.74** [0.69, 0.78] | **0.76** [0.70, 0.82] | **0.73** [0.66, 0.78] | **0.74** [0.69, 0.79] |
| DeepSeek-V3 | 0.30 | 0.70 [0.66, 0.74] | 0.66 [0.60, 0.72] | 0.72 [0.65, 0.78] | 0.69 [0.63, 0.74] |
| Claude Opus | 0.35 | 0.71 [0.66, 0.75] | 0.73 [0.67, 0.80] | 0.70 [0.63, 0.76] | 0.72 [0.66, 0.76] |
| GPT-5 | 0.25 | 0.67 [0.63, 0.72] | 0.58 [0.51, 0.65] | 0.71 [0.64, 0.78] | 0.64 [0.58, 0.70] |
| GPT-4o | 0.18 | 0.66 [0.61, 0.70] | 0.66 [0.60, 0.73] | 0.66 [0.59, 0.72] | 0.66 [0.60, 0.71] |
| GPT-oss-120B | 0.23 | 0.66 [0.61, 0.70] | 0.59 [0.53, 0.66] | 0.68 [0.61, 0.75] | 0.63 [0.57, 0.69] |
| Qwen3-32B | 0.30 | 0.65 [0.60, 0.69] | 0.51 [0.44, 0.57] | 0.71 [0.63, 0.78] | 0.59 [0.53, 0.65] |
| Gemma3-27B | 0.25 | 0.64 [0.59, 0.68] | 0.75 [0.70, 0.81] | 0.61 [0.55, 0.67] | 0.67 [0.63, 0.72] |
| Qwen3-4B | 0.07 | 0.65 [0.60, 0.69] | 0.62 [0.56, 0.69] | 0.65 [0.59, 0.72] | 0.64 [0.58, 0.69] |

Thresholds chosen by maximizing Youden’s index on the case-level bootstrap.
Values in brackets indicate 95% confidence intervals.
Bold indicates the best-performing model for each metric.

CLMBR-T showed modest discrimination improvement over physicians with an IDI of 0.19 (95% CI: 0.13-0.25), indicating enhanced ability to separate patients who experienced adverse outcomes from those who did not. In contrast, other models showed minimal improvements with confidence intervals spanning zero (Claude: 0.02 [-0.02, 0.06]; Deepseek: 0.009 [-0.03, 0.05]; GPT-5: 0.02 [-0.02, 0.06]). Systematic differences in risk scale utilization were evident: physicians used 2.0-5.0, CLMBR-T employed the full 1.0-5.0 range, Claude operated within a compressed 2.0-4.0 range, Deepseek used exclusively moderate-to-high ratings (3.0-5.0), and GPT-5 spanned 2.0-5.0. These scale differences produced conflicting reclassification metrics, with CLMBR-T showing large positive continuous NRI (0.39) but near-zero categorical NRI (−0.03), reflecting systematic calibration artifacts rather than meaningful clinical reclassification improvements.

### 3.5 Confidence impact and human alignment

The relationship between self-reported confidence and predictive accuracy varied markedly across methods (Table 7). CLMBR-T demonstrated the strongest confidence calibration with a significant confidence-performance correlation (r=0.212, p*<*0.001) and clear performance stratification from 67% accuracy in low-confidence predictions to 87% accuracy in high-confidence predictions. Notably, CLMBR-T exhibited systematic underconfidence—even its low-confidence predictions achieved reasonable accuracy, suggesting the model remained reliable when expressing uncertainty.

**Table 7:**
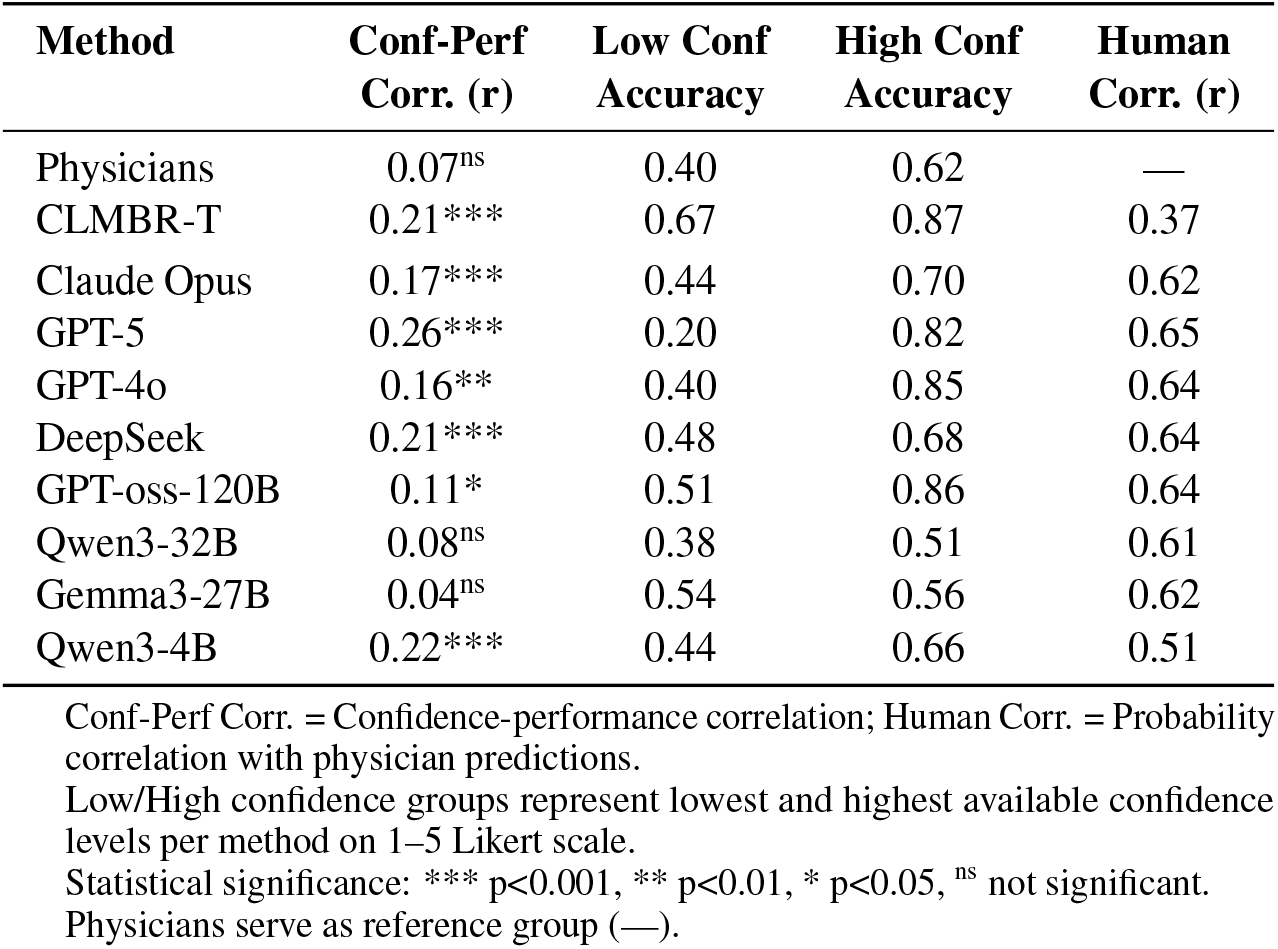
Confidence calibration, human alignment and performance stratification across methods.

In contrast, physicians showed poor confidence calibration with a weak, non-significant confidence-performance correlation (r=0.068, p=0.173). Doctors were systematically overconfident, with high confidence ratings failing to predict better performance.

LLMs showed mixed confidence patterns. Most models showed significant confidence-performance relationships, with GPT-5 exhibiting the largest confidence-performance gap (+62% accuracy improvement from low to high confidence). However, most LLMs were generally overconfident, with the notable exceptions of Claude Opus and GPT-oss-120B, which achieved well-calibrated confidence ratings.

Analysis of human reasoning alignment revealed that LLMs better captured human decision-making patterns than CLMBR-T (Table 7). Taking into account probability correlations, Claude 4.1 Opus showed the strongest alignment with human reasoning, achieving the highest probability correlation of 0.618 with physician predictions and well-calibrated confidence patterns. Other large LLMs (GPT-5, GPT-4o, DeepSeek) also showed strong human alignment, with probability correlations ranging from 0.64 to 0.65. CLMBR-T exhibited the poorest alignment with human reasoning despite its superior discriminative performance, showing the weakest probability correlation with physicians (r=0.370).

## 4 Discussion

This study shows that when physicians, structured ML models, and off-the-shelf LLMs are evaluated under identical data conditions, their prognostic performance differs in systematic and clinically meaningful ways. Prior work has characterized physician prognostic accuracy [4, 8] and compared clinicians with predictive models in settings such as hospital readmissions and ED visits [5, 9, 10, 11], but no previous study has directly contrasted physicians, state-of-the-art LLMs, and structured ML models using a controlled, head-to-head design. By holding information constant across evaluators, this work isolates the intrinsic strengths and weaknesses of human versus algorithmic forecasting, offering new insights into not only relative accuracy but also alignment with clinical reasoning and confidence calibration.

The results of this study show a clear performance hierarchy: CLMBR-T achieved superior discriminative performance (AUROC 0.793, 95% CI: 0.745–0.833) compared to pooled physicians (AUROC 0.65, 95% CI: 0.59–0.70), while all LLMs—from large models like GPT-5 and Claude Opus 4.1 (AUROC *∼* 0.74) to smaller variants like Qwen3-4B (AUROC 0.67)—performed at intermediate levels. This pattern held across both prediction endpoints and classification metrics, establishing that structured ML models consistently outperform human clinical judgment for this forecasting task.These findings align with prior literature showing that predictive models generally achieve modestly higher discrimination and better calibration than unaided physicians in prognostic settings [11, 12]. At the same time, the results extend previous work by showing this gap under head-to-head conditions with contemporary LLMs, while also reaffirming decades of evidence that physician prognostic estimates are often inaccurate, overly optimistic, and highly variable [4, 8].

Beyond accuracy, these findings reveal a trade-off between predictive performance and alignment with human reasoning. While CLMBR-T achieved the highest accuracy, it exhibited the weakest alignment with physician decision-making patterns (correlation *r* = 0.37). In contrast, LLMs showed stronger correlations with physician reasoning (*r* = 0.51– 0.65), in some cases matching or exceeding typical inter-physician agreement levels (*r* = 0.56). This echoes prior observations that structured models frequently identify non-intuitive patterns that diverge from human reasoning [13], while LLMs, trained on human-generated text, naturally approximate cognitive heuristics and medical gestalt. Such alignment may facilitate clinical adoption even when pure predictive performance is lower. Similar reasoning has been proposed in comparative studies where physician judgment was found to outperform models in more complex, less standardized care settings, such as critical illness [11], underscoring the importance of balancing accuracy with interpretability.

Marked differences emerged in confidence calibration, with implications for safe adoption. Physicians demonstrated poor calibration, showing systematic overconfidence and a weak relationship between certainty and accuracy (*r* = 0.07, *p* = 0.17). This aligns with extensive literature documenting that physician confidence is only weakly correlated with actual accuracy and is often inflated by optimism and other cognitive biases [14, 15, 16]. By contrast, CLMBR-T exhibited the strongest confidence-performance relationship (*r* = 0.21, *p <* 0.001) but was systematically underconfident. Such conservatism may paradoxically enhance safety, as it reduces the risk of automation bias and unwarranted trust. Among LLMs, most demonstrated significant confidence-performance correlations, suggesting a degree of “self-awareness” about prediction reliability. GPT-5 demonstrated the strongest such relationship (*r* = 0.26, *p <* 0.001), followed by Qwen3-4B, DeepSeek, and Claude Opus. While some overconfidence persisted, Claude Opus and GPT-oss-120B achieved good overall calibration, approximating an optimal compromise between accuracy and interpretability. The ability of LLMs to provide both moderate accuracy and trustworthy uncertainty estimates contrasts sharply with physicians’ poor calibration, reinforcing prior findings that unaided clinical judgment is poorly aligned with actual risk [8, 9].

The substantial variability among individual physicians (AUROC range: 0.55–0.83) highlights the heterogeneity of clinical reasoning. Three participants exceeded CLMBR-T’s benchmark, while others performed substantially worse. Pooled averages therefore obscure the presence of both high- and low-performing clinicians. We can observe a trend toward better prognostic performance among physicians with 5–10 years of experience, consistent with prior studies suggesting that mid-career physicians may achieve peak forecasting ability [5, **?**]. This effect may reflect the influence of accumulated experience, particularly in emergency and acute care contexts where exposure to patient outcomes calibrates prognostic intuition [17]. In contrast, younger physicians and senior physicians who are less engaged in ED work may lack this calibration.

These findings suggest that hybrid approaches could harness complementary strengths. CLMBR-T could serve as a high-performance screening tool, flagging high-risk patients with conservative probability estimates. LLMs, particularly those with strong reasoning alignment and calibrated confidence, could then provide interpretable rationales and context-sensitive explanations to clinicians. Such layered designs may enhance both predictive accuracy and clinical acceptance, echoing previous evidence that combined human–machine approaches often outperform either approach alone [18, 19].

Several limitations warrant consideration. First, correlation was assessed with human predictions rather than true reasoning alignment; probability correlations provide insight into decision-making patterns but not underlying cognitive processes. Second, Likert-scale physician ratings compared against probability outputs may have introduced scaling biases, despite sensitivity testing. Third, synthetic vignettes—while carefully constructed—cannot fully capture the complexity of real-world encounters, where clinicians draw on contextual and institutional knowledge. This constraint may have disadvantaged physicians relative to models. The relatively small sample size (35 physicians from a single healthcare system) further limits generalizability. Findings may not extrapolate to broader clinician populations or practice contexts. The wide variability in physician performance also underscores the limitations of pooled “average clinician” comparisons, suggesting that individualized benchmarks or adaptive support systems may be more appropriate [10, 12]. Finally, results are restricted to one prognostic task within a single institutional context, and the rapid evolution of LLMs capabilities limits temporal validity. Beyond technical considerations, regulatory and ethical implications—including safety, accountability, and transparency—must be addressed before such systems can be responsibly deployed in clinical care.

## 5 Conclusion

This head-to-head comparison establishes that ML models trained on structured EHR data outperform both physicians and LLMs in predictive accuracy and confidence calibration for forecasting unplanned hospital admissions, though LLMs achieved competitive zero-shot performance and better approximated human clinical reasoning patterns. The substantial variability in physician performance, with three out of 35 physicians exceeding the ML benchmark, highlights limitations of benchmarking against “average” clinical judgment. These findings suggest that hybrid approaches combining high-performance ML screening with interpretable LLM explanations may optimize both accuracy and clinical adoption, addressing the fundamental trade-off between algorithmic performance and human reasoning alignment that characterizes artificial intelligence implementation in healthcare settings.

## Data Availability

All data produced in the present study are available upon reasonable request to the authors

